# Who is responsible for nurse wellbeing in a crisis? A single centre perspective

**DOI:** 10.1101/2023.03.29.23287904

**Authors:** Luke Hughes, Anika Petrella, Lorna A Fern, Rachel M Taylor

## Abstract

**Background:** Leadership during the COVID-19 pandemic often manifested as a command-and-control style of leadership which had detrimental emotional impacts on staff particularly the nursing workforce. Organisational responsibility for staff wellbeing would be necessary in another pandemic and leadership emerged as a key indicator of the overall health of an organisation and its workforce. Leadership can have detrimental effects on staff wellbeing or it can greatly boost their ability to handle a crisis. We sought to explore the interrelationship between leadership and nurses’ wellbeing in an inner-city university hospital during the initial wave of the pandemic.

**Methods:** Secondary analysis of interview data collected during a hospital-wide evaluation of barriers and facilitators to changes implemented to support the surge of COVID-19 related admissions during wave 1. Data were collected through semi-structured video interviews between May and July 2020. Interviews were analysed using Framework analysis

**Results:** Thirty-one nurses participated including matrons (n=7), sisters (n=8) and specialist nursing roles (n=16). Three overarching themes were identified: impact on nurses, personal factors and organisational factors. The impact on nurses manifested as distress and fatigue. Coping and help-seeking behaviours were found to be the two personal factors which underpinned nurses’ wellbeing. The organisational factors that impacted nurses’ wellbeing included decision-making, duty and teamwork.

**Conclusions:** The wellbeing of the workforce is pivotal to the health service, and it is mutually beneficial for patients, staff, and leaders. Addressing how beliefs and misconceptions around wellbeing are communicated, and accessing psychological support is a key priority to support nurses during pandemics.

## Introduction

The physical and emotional burden of the nursing role is internationally recognised^1^. While these tribulations are accepted elements of the role, global pandemics place nurses in a uniquely vulnerable position particularly in healthcare settings where resources are already under significant pressure due to staffing and financial shortages^2-4^. The additional emotional distress placed on nurses during a pandemic can increases burnout and disengagement, and this has shown to persist following control or cessation of the pandemic^3,5^. A key factor associated with burnout and disengagement is additional work pressures such as redeployment in response to pandemics. Previous pandemics have shown that redeployment is essential in responding to a crisis, however it was also associated with poorer wellbeing during the Severe Acute Respiratory (SARS) pandemic and therefore was predicted to impact nurses during the current COVID19 pandemic^6,7^. Additional concerns for the impact on nursing welfare was the ease of spread and transmission of COVID19, where SARS was primarily a nosocomial infection with limited community transmission, COVID-19 is a disease characterised by high pathogenicity and virulence^8^. Stressors which were identified during SARS (e.g., fear of transmission to family) were therefore going to be significantly more present for nurses during the early phases of COVID^3,7^. Due to the much larger scale outbreak of COVID-19 and its associated rates of mortality, nurses were predicted to be at a high risk of long-lasting distress and systemic burnout without intervention^4^. It was anticipated this would be on a much wider scale than SARS, given the global impact of the virus of which the likes has not been witnessed in recent history. The United Kingdom (UK) observed the pandemic developing in other countries, and the situation in Italy was a source of concentrated anxiety and fear among healthcare professionals already anticipating and bracing themselves for its arrival^9^. Of concern, was the insight into the detrimental and lasting impact on previous pandemics on the nursing workforce. The question asked by many was how would the National Health Service (NHS) protect its workforce?

Evidence shows that redeployment, social isolation and poor cohesion within teams are risk factors to nurse wellbeing during a pandemic^7^. Traditionally, healthcare services have focused on the ways staff ‘cope’ in times of crisis in order to remain resilient. In literature, coping is traditionally thought of as a dynamic process that promotes survival and adaption as a response to stimuli which are perceived as threatening^10^. These multifaceted responses are fuelled by learned patterns of ideas and behaviours influenced by personality traits, historical relationships and situational stressors^11^. For example, during SARS adaptive coping mechanisms such as positive reframing was observed to support nurses’ wellbeing, whereas more maladaptive coping responses, such as avoidance or self-blame, led to poorer outcomes^7^. Previous research has shown that nurses who employed more maladaptive coping styles were more vulnerable to burnout^12^. However, the literature around coping has been criticised for being vague and inconsistent^13^. This has the undesired effect of creating feelings of self-blame and a feeling of being unsupported; where in reality, resilience should be thought of as a collective and organisational responsibility^1^. Despite this, studies often recommend creating a work environment which promotes healthy and adaptable coping mechanisms in response to crises such as COVID-19 in order to maintain resilience. It was clear from the research on SARS that the way the healthcare system dealt with crisis was necessary to avoid further undue harm – but in practicality what this would look like lacked realistic structure.

Emerging evidence shows that COVID-19 highlighted the precarity of healthcare systems worldwide, and the vulnerability of its workforce^14^. In the wake of the pandemic emphasis was placed on the need for the visibility of leadership^3, 15^. Commentaries have hinged on the personal responsibility of the individual nurse to have the coping mechanisms and resilience to survive the trauma inherent to their duty. But is the responsibility also on the organisation and healthcare system as a whole to protect their staff? Placing this burden on staff who are already emotionally depleted is likely to result in compassion fatigue, which is likely to affect the standard of care staff are able to provide^14^. We have previously shown the importance of nursing leadership during the pandemic and the way the command-and-control style of leadership impacted on nurses at each level of the hierarchy^16^. At the heart of this was the subsequent emotional impact. There have been calls for more compassionate leadership within organisations^14^. This study focuses on the impact of leadership on the wellbeing of nurses working in an inner-city university hospital during the initial wave of the COVID19 pandemic.

## Methods

### Participants

The evaluation was conducted in accordance with the UK Framework for Health and Social Care Research (Health Research Authority (HRA)^17^. The HRA has the Research Ethics Service as one of its core functions and they determined the evaluation these data were originally obtained for was exempt from the need to obtain approval from an NHS Research Ethics Committee. A convenience sample of nurses at different levels of seniority, participants were invited through the Trusts group email lists. The purpose of the evaluation was explained to participants at the beginning of the video call, who then were given the opportunity to ask questions. If they were happy to continue, they were asked to give a recorded consent. All participants were able to stop the interview at any time and were assured of anonymity and confidentiality. Interviews were transcribed and anonymised. Voice recordings were deleted, and the anonymised transcripts were stored on a password protected NHS computer system. Approval for secondary analysis was provided by the hospital head of research governance.

### Data collection

Data were collected through individual semi-structured interviews between May and July 2020 during and immediately following the first wave of COVID19. The guide for the interviews asked about elements of service change, which included a section on staff wellbeing and a section on emotional impact. Interviews were conducted through video conference software and were digitally transcribed verbatim. Interviews were performed by three researchers with experience of interviewing participants and providing emotional support so immediate support could be provided if nurses became distressed.

### Data analysis

Data were analysed using Framework Analysis^18^. Framework analysis enabled multiple researchers to review the coding to check for accuracy of the interpretation, and in doing so ensure the audibility of the findings. The Framework for the evaluation was developed from previous interviews with operational leads in the hospital setting. Impact on wellbeing and the emotional impact of the pandemic were identified as an overarching theme, underpinning every aspect of working in the pandemic and therefore a secondary framework was developed to focus specifically on this in order to further understand the emotional impact of nurse’s experiences. The main framework was developed by two members of the evaluation team, checked by an independent researcher with expertise in qualitative research; the secondary framework was reviewed by a third member of the evaluation team.

### Findings

A total of 31 nurses participated in the evaluation. This included matrons (n=7), sisters (n=8) and nurses in specialist roles (n=16). Interviews were 40-60 minutes in length. The findings of the study were divided into three major themes: impact on nurses, personal factors and organisational factors. Supportive quotes are presented in Table 1.

**Table 1:**
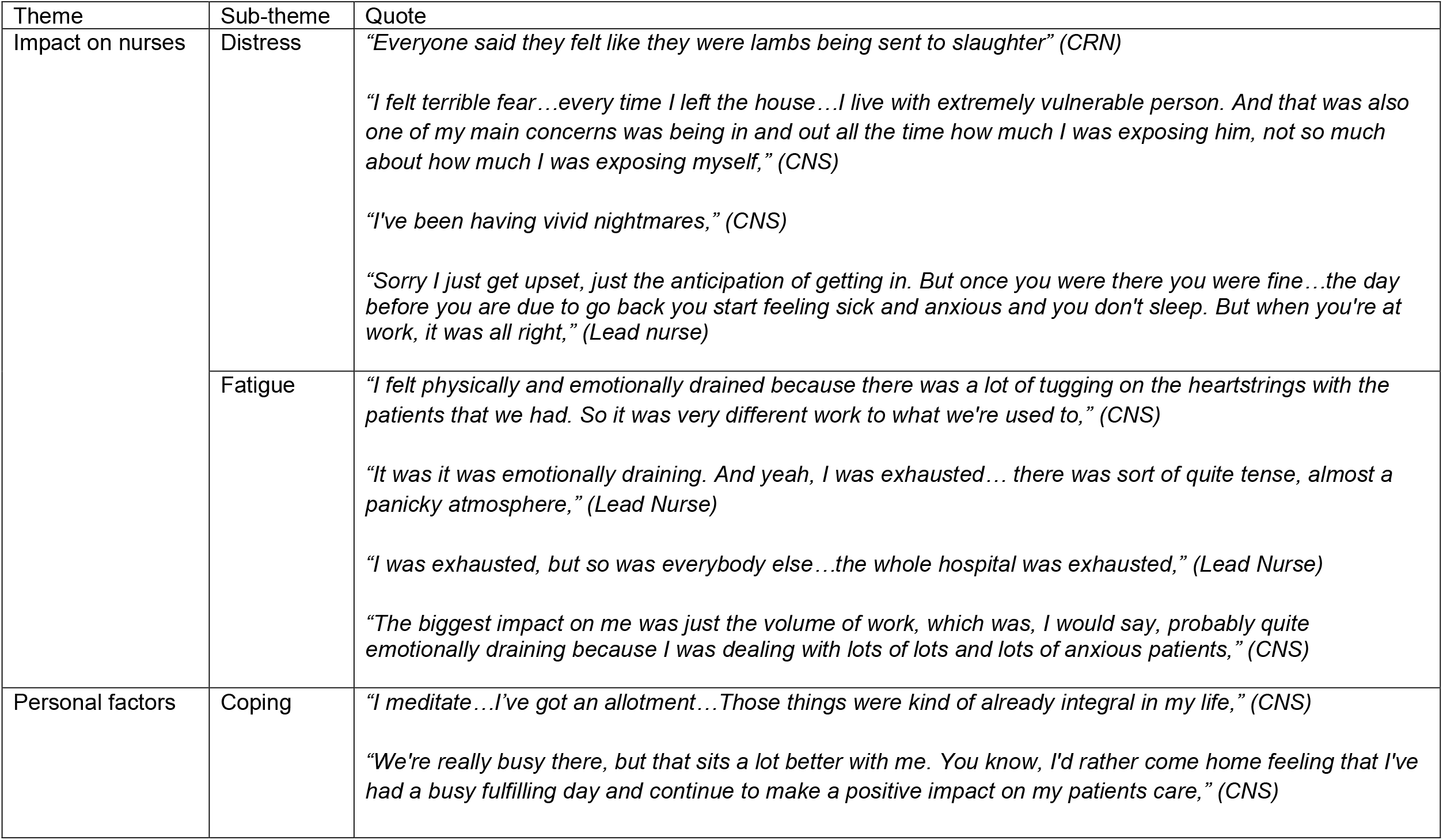
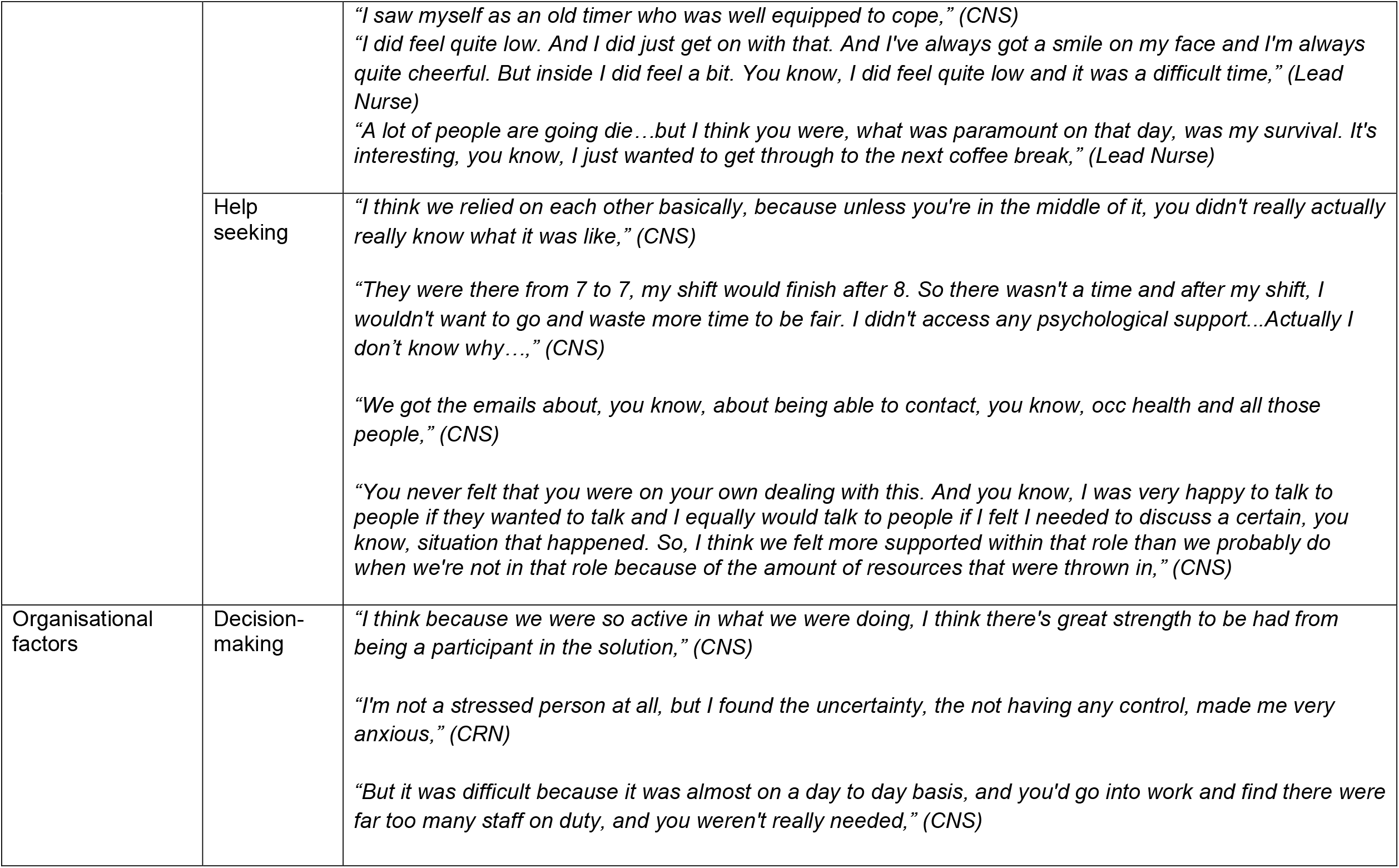
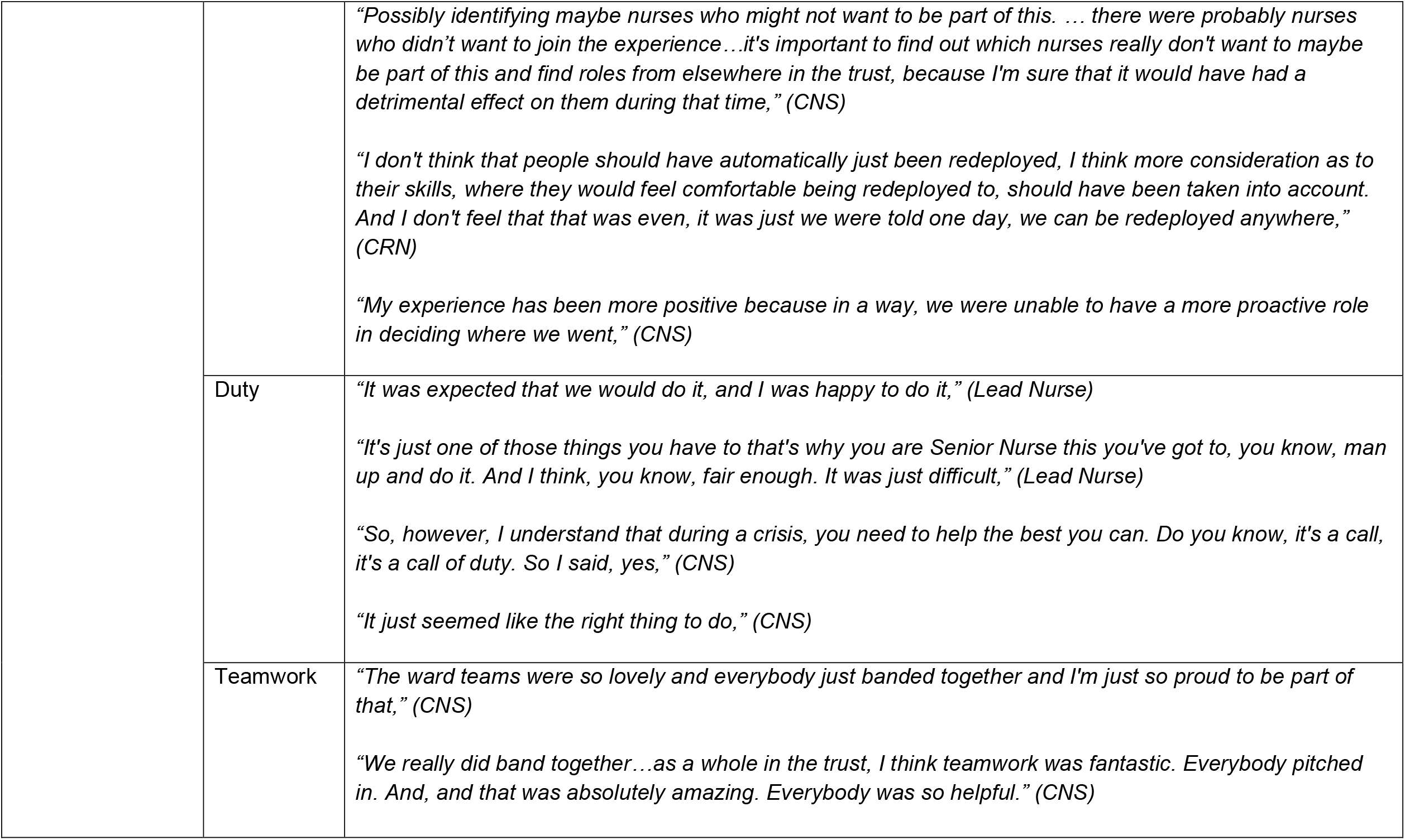
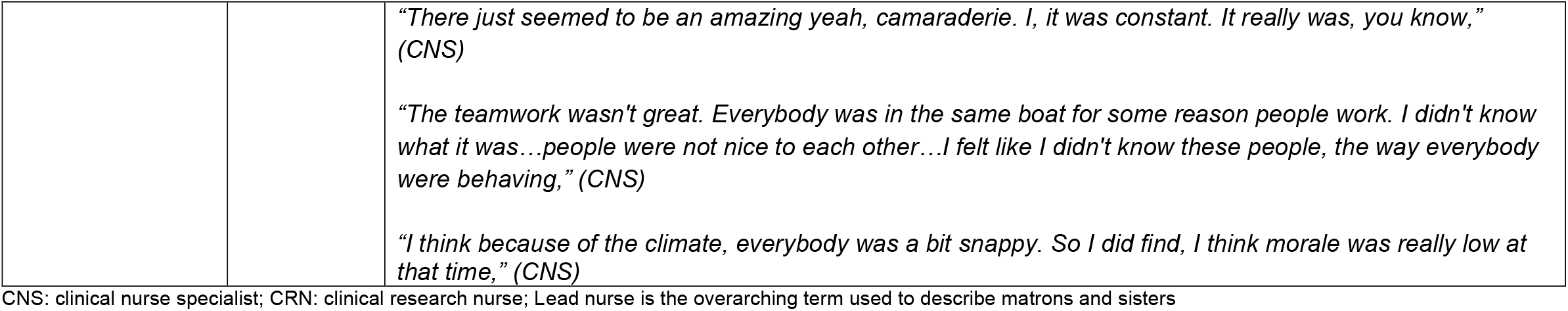
Supporting quotes

| Theme | Sub-theme | Quote |
| --- | --- | --- |
| Impact on nurses | Distress | *"Everyone said they felt like they were lambs being sent to slaughter" (CRN)*<br>*"I felt terrible fear...every time I left the house...I live with extremely vulnerable person. And that was also one of my main concerns was being in and out all the time how much I was exposing him, not so much about how much I was exposing myself," (CNS)*<br>*"I've been having vivid nightmares," (CNS)*<br>*"Sorry I just get upset, just the anticipation of getting in. But once you were there you were fine...the day before you are due to go back you start feeling sick and anxious and you don't sleep. But when you're at work, it was all right," (Lead nurse)* |
|  | Fatigue | *"I felt physically and emotionally drained because there was a lot of tugging on the heartstrings with the patients that we had. So it was very different work to what we're used to," (CNS)*<br>*"It was it was emotionally draining. And yeah, I was exhausted... there was sort of quite tense, almost a panicky atmosphere," (Lead Nurse)*<br>*"I was exhausted, but so was everybody else...the whole hospital was exhausted," (Lead Nurse)*<br>*"The biggest impact on me was just the volume of work, which was, I would say, probably quite emotionally draining because I was dealing with lots of lots and lots of anxious patients," (CNS)* |
| Personal factors | Coping | *"I meditate...I've got an allotment...Those things were kind of already integral in my life," (CNS)*<br>*"We're really busy there, but that sits a lot better with me. You know, I'd rather come home feeling that I've had a busy fulfilling day and continue to make a positive impact on my patients care," (CNS)* |
|  |  | *"I saw myself as an old timer who was well equipped to cope," (CNS)*<br>*"I did feel quite low. And I did just get on with that. And I've always got a smile on my face and I'm always quite cheerful. But inside I did feel a bit. You know, I did feel quite low and it was a difficult time," (Lead Nurse)*<br>*"A lot of people are going die...but I think you were, what was paramount on that day, was my survival. It's interesting, you know, I just wanted to get through to the next coffee break," (Lead Nurse)* |
|  | Help seeking | *"I think we relied on each other basically, because unless you're in the middle of it, you didn't really actually really know what it was like," (CNS)*<br>*"They were there from 7 to 7, my shift would finish after 8. So there wasn't a time and after my shift, I wouldn't want to go and waste more time to be fair. I didn't access any psychological support...Actually I don't know why...", (CNS)*<br>*"We got the emails about, you know, about being able to contact, you know, occ health and all those people," (CNS)*<br>*"You never felt that you were on your own dealing with this. And you know, I was very happy to talk to people if they wanted to talk and I equally would talk to people if I felt I needed to discuss a certain, you know, situation that happened. So, I think we felt more supported within that role than we probably do when we're not in that role because of the amount of resources that were thrown in," (CNS)* |
| Organisational factors | Decision-making | *"I think because we were so active in what we were doing, I think there's great strength to be had from being a participant in the solution," (CNS)*<br>*"I'm not a stressed person at all, but I found the uncertainty, the not having any control, made me very anxious," (CRN)*<br>*"But it was difficult because it was almost on a day to day basis, and you'd go into work and find there were far too many staff on duty, and you weren't really needed," (CNS)* |
|  |  | *"Possibly identifying maybe nurses who might not want to be part of this. ... there were probably nurses who didn't want to join the experience...it's important to find out which nurses really don't want to maybe be part of this and find roles from elsewhere in the trust, because I'm sure that it would have had a detrimental effect on them during that time," (CNS)*<br>*"I don't think that people should have automatically just been redeployed, I think more consideration as to their skills, where they would feel comfortable being redeployed to, should have been taken into account. And I don't feel that that was even, it was just we were told one day, we can be redeployed anywhere," (CRN)*<br>*"My experience has been more positive because in a way, we were unable to have a more proactive role in deciding where we went," (CNS)* |
|  | Duty | *"It was expected that we would do it, and I was happy to do it," (Lead Nurse)*<br>*"It's just one of those things you have to that's why you are Senior Nurse this you've got to, you know, man up and do it. And I think, you know, fair enough. It was just difficult," (Lead Nurse)*<br>*"So, however, I understand that during a crisis, you need to help the best you can. Do you know, it's a call, it's a call of duty. So I said, yes," (CNS)*<br>*"It just seemed like the right thing to do," (CNS)* |
|  | Teamwork | *"The ward teams were so lovely and everybody just banded together and I'm just so proud to be part of that," (CNS)*<br>*"We really did band together...as a whole in the trust, I think teamwork was fantastic. Everybody pitched in. And, and that was absolutely amazing. Everybody was so helpful." (CNS)* |
|  |  | *"There just seemed to be an amazing yeah, camaraderie. I, it was constant. It really was, you know," (CNS)*<br>*"The teamwork wasn't great. Everybody was in the same boat for some reason people work. I didn't know what it was...people were not nice to each other...I felt like I didn't know these people, the way everybody were behaving," (CNS)*<br>*"I think because of the climate, everybody was a bit snappy. So I did find, I think morale was really low at that time," (CNS)* |

CNS: clinical nurse specialist; CRN: clinical research nurse; Lead nurse is the overarching term used to describe matrons and sisters

### Impact on nurses

Impact on nurses manifested as distress and fatigue.

Distress was common amongst participants. At the forefront of this distress was the unknown nature of the virus. The uncertainty of the situation was destabilising and created anxiety in the early stages of the pandemic. This largely revolved around the NHS rapidly adjusting to engage with the virus, with many staff not feeling safe or protected from transmission. Many nurses noted they experienced anxiety around bringing the infection home to their family, particularly if they had children or lived with a vulnerable person. Those who were vulnerable themselves were also understandably frightened. Nurses expressed concerns over the inconsistent rules in those early months, particularly around depleted stock of adequate personal protective equipment (PPE), which left many feeling nervous and unsure whilst on the frontline. There were a few nurses who had very difficult experiences during their redeployment. This resulted in some reconsidering their roles in healthcare.

Nurses reported poor emotional wellbeing, experiencing nightmares, flashbacks and other manifestations of traumatic stress following the end of the first wave, and felt hurt and unsupported by the system they expected to protect them. However, other nurses said that despite the high level of distress the pandemic had elicited, they would still be likely to return to those working conditions if needed. Some were more ambivalent about this, and a small number disclosed that they would not be able to return to work if they were redeployed in the same manner again.

Participants expressed both physical and emotional fatigue, as well as disillusionment with their work. The long work hours were exhausting, particularly when combined with working in PPE. Overall, the nature of COVID-19 resulted in a high degree of emotionally draining work for nurses, due to the high number of critically ill patients and the restrictions on visitations. Many patients were admitted and subsequently died alone, which nurses found very hard to witness. Many spoke about trying to support the emotional wellbeing of their patients on the wards, but the pressures of their workloads interfered with them being as present as they would normally be for terminal patients. This in turn caused conflict and guilt.

### Personal Factors

Coping and help-seeking behaviours were found to be the two personal factors underpinning nurses’ wellbeing.

Those in more senior roles felt they had good coping mechanisms which were established prior to the pandemic. These mechanisms were how they normally dealt with their occupational stressors, which initially helped them to process distress as they faced the unknown threat of the pandemic. These coping mechanisms were varied from adaptive engagement such as gardening, crafting or exercising, to emotion focused and social coping, such as speaking about their emotions and experiences to co-workers, friends and family. Speaking to colleagues was shown to be a form of coping that was highly advocated by most nurses. This was important in providing stability during difficult times on the frontline. Finding a sense of meaning in their work was believed to be protective, many feeling pride and accomplishment.

Many nurses expressed feeling confident and resilient when it came to dealing with emotional stresses related to their work. In particular staff who were more senior felt that they were more experienced handling these intense situations. In turn they spoke about using their resilience to help protect the rest of their team. Equally, nurses discussed feeling they had been able to adapt their ways of working to manage the intensity in workload. While many had felt apprehensive before being redeployed some found the challenge invigorating. However, some staff noted that their normal forms of coping were not always effective at combating the distress inherent to COVID-19. For example, some found their normal outlets were simply overwhelmed by the pressures brought by the pandemic, while others had normal avenues for coping removed due to government social restrictions, such as attending religious gatherings or going to the gym. Some participants engaged in unhelpful forms of coping such as avoidance and self-blame.

Throughout COVID-19 the hospital diverted psychology resources to the staff psychological services and occupational health teams, yet the view from many nurses was that they actually preferred to get their emotional support from colleagues. Many noted they did not have the time to access mental health support services, and while they were still in the midst of the crisis they did not think it would be effective. There was also a reluctance to engage with staff psychology services as they felt they would not understand their experiences to the extent their colleagues would. For those who had experienced distressing circumstances their views were mixed. Some sought out psychological support, either through the hospital or privately. Some however continued to be reluctant to engage with psychological support, despite expressing unresolved emotional distress. These nurses expressed elements of avoidant coping in their thought processes, by claiming they did not have time or that psychology would not be beneficial. Some did not know how to engage with services and what supports were available while others expressed that they did not feel those services would be useful for them.

### Organisational Factors

The organisational factors that impacted nurses’ wellbeing included decision-making, duty and teamwork.

The decision to redeploy a nurse without any consultation had a visible impact. Where nurses felt that they had been consulted about being redeployed and that their professional skill set and personal circumstances were considered, they were more likely to express a sense of confidence about the experience. Nurses who expressed feeling a sense of agency tended to frame the pandemic as a matter of their professional duty and described feeling ready and willing to go where they were needed. They also tended to better employ problem-solving behaviours. Nurses who felt they had no say in the process and described management as not taking their skills or personal circumstances into account felt anxious and distressed about redeployment. Some nurses described having little to no conversations with management other than being told where they would be moved, which led to frustration and apprehension. Many in this situation felt management were not thinking about nurses as people and found this more authoritarian approach destabilising. Feeling powerless in the face of redeployment left some to catastrophise. Even nurses who had personal feelings of control and agency recognised that not all their colleagues felt the same way. It was suggested that a system was needed to screen staff to identify those willing to be redeployed, their relevant skills and where they would be willing to go. While some reflected that this might not have been possible in the limited time the hospital had to prepare, others were less understanding and questioned why hospital leaders had not forethought the need for this already. Regardless, everyone felt that redeployment should be coordinated uniformly for all nurses.

For nurse leaders, the lack of control was doubly frustrating. Some felt as though they were perpetually caught between the decisions of higher management, and the worries and concerns of their own staff. This was more so at the beginning of the pandemic, when rules around PPE changed frequently; they felt as though they were giving their staff inconsistent support and worried they were unknowingly exposing their staff to risks. Furthermore they felt as though they were often the bearers of bad news which impacted their relationships with their staff. Some expressed frustration with senior management who made decisions which came into conflict with their own choices they had made with their staff’s personal circumstances in mind.

Many nurses’ felt that despite their own worries or fears it was their responsibility to do what was needed during a time of crisis. For some, this made the choice simpler; they went where they were needed. This was particularly common among more senior nurses who expressed feeling conflict with their sense of duty and their own anxieties. They expressed feeling frightened of working on COVID-19 wards but knew it was their role to lead by example.

Despite this they felt anxious working on the frontline and honour bound to put on a brave face despite their own fears.

Teamwork was a core element which helped nurses endure this experience. They noted the importance of teamwork in the everyday life of their roles being magnified in the experience of redeployment, and how teams banded together to support each other through this challenging time. It was felt that staff were more likely than ever to help each other and support one another. Some felt the experience of redeployment flattened the hierarchy common in medical wards and allowed for staff of all levels to work together side-by-side. Structured management of these teams was an important aspect of their success. When teams were constructed and led with care and attentiveness, the sense of camaraderie was palpable. Teams who experienced strong visible leadership were better able to maintain morale. This camaraderie helped to foster a sense of community and meaningfulness in the work they were doing.

Making sure that host teams were receptive and welcoming was underpinned by a clear sense of leadership. However, there were some who had more negative experiences. Conflict seemed to arise particularly when nurses were redeployed into areas of high stress acuity. Host ward hostility led to nurses having negative experiences, feeling isolated from their team and struggling to cope. This tended to result from a lack of leadership, or staff not being directed to help others adapt to a new role. Whereas the camaraderie mentioned by others was protective, nurses who experienced unsupportive teams were more likely to be exposed to emotional distress. While some managed to establish an effective working system, there was a clear benefit from keeping teams together. Teams who stayed together maintained their previously built rapport and supported one another. Teams made of various new and disparate nurses from different backgrounds struggled to come together when lacking a clear sense of leadership.

## Discussion

The findings of this study reflect the literature of past pandemics and emerging studies around the experiences of nurses in the COVID-19 climate^2,3,7,15^. In addition to these findings, we have shown the importance of leadership in ameliorating some of the negative impacts on nurse wellbeing. It was already clear that nurses found their work taxing both physically and emotionally in pre-pandemic times, and that this stress was magnified greatly in the pandemic^2,7^. Consistent with the model published by the British Psychological Society (BPS), the initial stages were marked by heightened states of anxiety and stress, followed by a surge towards action and a pervading air of heroism^6^. The BPS warned that this surge to solution would result in a period of disillusionment if steps were not taken to protect staff wellbeing. While historically the task of coping would often fall to nurses themselves, the message from SARS was clear – failure to protect staff could land a blow to the workforce during a vulnerable period^2,14^. The findings of this study offer some perspective on how nurses experienced the initial wave of the pandemic and what aspects of leadership may have helped to protect them.

It came as no surprise that nurses spoke about experiences of emotional distress. For the most part, nurses had gone into the experience knowing they were about to be challenged and were thus ambivalently ready for COVID-19. However, this distress has become prolonged as the pandemic continues without a clear end in sight. Prolonged exposure to stressful stimuli should be of marked concern as it greatly increases the likelihood of staff experiencing affective and post-traumatic stress symptoms^7^. Fatigue was also frequently discussed which is another common outcome of repeated stress exposure, particularly with the pandemic^3,5^. Psychological distress is a leading factor contributing to burnout, a pattern frequently observed in SARS^7^. Unequivocally, the pandemic has greatly impacted nurses. Nurses who relied on previously established coping styles which were normally adaptive found that they were compromised and overwhelmed by the disruption that COVID-19 had brought^2^. During the pandemic, nurses felt conflicted by their duty of care and their very real human fears of a then little understood virus. Nurses were expected to be able to care for their patients, while also caring for themselves in the process. Failure to do so, by experiencing distress and fatigue, was more likely to illicit feelings of self-criticism and result in a lack of self-compassion^14^. This is particularly relevant within an intense work culture, where being perceived to lack resilience can have unwanted negative connotations^1^. Some participants spoke of the need to put on a *‘brave face’* which aligns narratively with the BPS prediction of ‘heroism’ and our own theme of duty. However normalising fear and anxiety and allowing space for accepting these feelings as being valid was more likely to result in nurses feeling able to cope. This acceptance and compassion to self is not the responsibility of the individual rather something which needs to be hardwired into organisational practices^14,19^. The cornerstone of this practise is compassionate leadership, rather than command-and-control style^14,16,20^.

The sample showed a reluctance to seek help. Nurses often relied on the support of their colleagues when experiencing emotional distress, and in this sample teamwork was an extremely vital meditator which protected many. However, many showed avoidant and dismissive attitudes towards psychological support, which is historically observed in the profession^10^. This may be a product of how services were offered, the type of services which were available, or point to ingrained institutional beliefs. It is difficult to say that nurses are not being supported by their organisations, when they are also not actively engaging in the support which is being offered^3^. Lastly, it was clear that decision making from leadership without dialogue with nurses was a source of distress. When it was felt that the reasoning behind upper management’s decisions were unclear, it left nurses feeling frustrated and powerless. Having some sense of control in difficult situations^19^. Decisions, such as redeploying some team member with little discussion or thought of their personal circumstances, greatly affected the wellbeing of nurses and was likely to cause them to express distress. Similarly breaking up teams, an important source of support, contributed highly to feelings of frustration and gave a message to some that management simply didn’t understand the experience of those on the frontline at all. Where nurses felt listened to it tended to reflect feeling positively supported and protected and valued by leadership.

The BPS outlined how strong visible leadership was an important factor in protecting staff^6^. Nurses who felt seen and supported by their managers were more likely to express an ability to adapt and cope with COVID-19, at least anecdotally. For those who felt they lacked leadership, or were redeployed to unwelcoming teams, the situation was framed much more negatively and morosely. Previously our research has shown how much an impact leadership across all levels can have on nurses, positively and negatively^16^. Similar themes surrounding the importance of team cohesion and reflective practice informing future protocols has been found in other samples of the COVID workforce as examples of strong leadership^15^. A culture of compassion needs to be threaded through leadership across all levels – this also included the need to protect and maintain the wellbeing of leaders themselves^14,15^. This suggests that perhaps it was the impact of organisational factors – rather than nurses own personal resilience - which played a pivotal role in their experience of the pandemic. In this regard, we might see coping styles and resilience not as static elements in a person’s character, rather as reactions to circumstances they inhabit. Compassionate leadership then emerges as a core mediator as to whether these strategies are likely to help or hinder the workforce.

## Conclusion

In a time where a controlled environment is difficult to maintain, and epistemic change happens at a rate hitherto unknown, it is critical to continue to understand the lived reality of those working on the frontline, and efficiently apply lessons learned from their experiences. Past evidence suggested that organisational responsibility for staff wellbeing would be necessary in the face of another pandemic. Recent reports show that there has been some increased effort to support staff both emotionally and practically during COVID-19^3^. It is positive to see these changes being made, and it is necessary to continue to learn from their experiences. Leadership emerged as a key indicator as to the overall health of an organisation and its workforce. Covid-19 has shown us that leadership can have detrimental effects on staff wellbeing or it can greatly boost their ability to handle a crisis^16^. The wellbeing of the workforce is pivotal to the health service, and it is mutually beneficial for patients, staff, and leaders^14^. Addressing beliefs and misconceptions around how messages around wellbeing are communicated and accessing psychological support is therefore another key priority. Perhaps most fundamental is the understanding that previous styles of authoritarian leadership during a crisis have not served to protect the workforce^20^. Moving forward in order to cultivate the natural resilience of the workforce, leadership needs to be grounded in compassionate practices which will only succeed when it is inbuilt into the organisation itself. To do this leadership should aim to provide a set of shared goals and vision with its workforce and reflect on past experiences to inform future practices. Approaches to wellbeing should be more flexible and dynamic, adapting to the needs and desires of the workforce rather than a generic approach^15^. Leaders must ground their work in understanding and advocacy for those they lead^20^.

## Data Availability

All data produced in the present work are contained in the manuscript

## Acknowledgements

We would like to thank the Nursing and Midwifery Leadership Team for commissioning and supporting the evaluation these data were derived from. We would also like to thank all the participants for giving us their time during the pandemic and for sharing so honestly about the impact of working in a pandemic.

## Contributors

RMT developed the protocol. LH, AP, LAF and RMT co-ordinated the running of the study, LH, AP, RMT were responsible for data acquisition. All the authors contributed to the analysis, drafted, critically revised and approved the final manuscript.

## Ethics approval

The data used in this study were generated from a project was assessed as service evaluation according to the toolkit published by the English Health Research Authority (HRA). As such, no formal research ethics approval was required. However, all participants consented in according to the UK Framework for Health and Social Care Research.

